# Interplay between the Genetics of Personality Traits, severe Psychiatric Disorders, and COVID-19 Host Genetics in the Susceptibility to SARS-CoV-2 Infection

**DOI:** 10.1101/2021.09.12.21263447

**Authors:** Urs Heilbronner, Fabian Streit, Thomas Vogl, Fanny Senner, Sabrina K. Schaupp, Daniela Reich-Erkelenz, Sergi Papiol, Mojtaba Oraki Kohshour, Farahnaz Klöhn-Saghatolislam, Janos L. Kalman, Maria Heilbronner, Katrin Gade, Ashley L. Comes, Monika Budde, Till F. M. Andlauer, Heike Anderson-Schmidt, Kristina Adorjan, Til Stürmer, Adrian Loerbroks, Manfred Amelang, Eric Poisel, Jerome Foo, Stefanie Heilmann-Heimbach, Andreas J. Forstner, Franziska Degenhardt, Jörg Zimmermann, Jens Wiltfang, Martin von Hagen, Carsten Spitzer, Max Schmauss, Eva Reininghaus, Jens Reimer, Carsten Konrad, Georg Juckel, Fabian U. Lang, Markus Jäger, Christian Figge, Andreas J. Fallgatter, Detlef E. Dietrich, Udo Dannlowski, Bernhardt T. Baune, Volker Arolt, Ion-George Anghelescu, Markus M. Nöthen, Stephanie H. Witt, Ole A. Andreassen, Chi-Hua Chen, Peter Falkai, Marcella Rietschel, Thomas G. Schulze, Eva C. Schulte

## Abstract

**Background:** The SARS-CoV-2 pandemic, with all its impacts on our way of life, is affecting our experiences and mental health. Notably, individuals with mental disorders have been reported to have a higher risk of contracting SARS-CoV-2. Personality traits could represent an important determinant of preventative health behavior and, therefore, the risk of contracting the virus.

**Aims:** We examined overlapping genetic underpinnings between major psychiatric disorders, personality traits, and susceptibility to SARS-CoV-2 infection.

**Methods:** Linkage disequilibrium score regression was used to explore the genetic correlations of COVID-19 susceptibility with psychiatric disorders and personality traits based on data from the largest available respective genome-wide association studies (GWAS). In two cohorts (the PsyCourse (n=1346) and the HeiDE (n=3266) study), polygenic risk scores were used to analyze if a genetic association between, psychiatric disorders, personality traits, and COVID-19 susceptibility exists in individual-level data.

**Results:** We observed no significant genetic correlations of COVID-19 susceptibility with psychiatric disorders. For personality traits, there was a significant genetic correlation for COVID-19 susceptibility with extraversion (p=1.47×10-5; rg=0.284). Yet, this was not reflected in individual-level data from the PsyCourse and HeiDE studies.

**Conclusions:** We identified no significant correlation between genetic risk factors for severe psychiatric disorders and genetic risk for COVID-19 susceptibility. Among the personality traits, extraversion showed evidence for a positive genetic association with COVID-19 susceptibility, in one but not in another setting. Overall, these findings highlight a complex contribution of genetic and non-genetic components in the interaction between COVID-19 susceptibility and personality traits or mental disorders.

## INTRODUCTION

The global spread of SARS-CoV-2 has revealed differences in susceptibility to and severity of SARS-CoV-2 infection at both the individual and the community level. Studies from different regions of the world suggest a rise in the incidence of psychiatric disorders due to the threat of the virus and the socioeconomic repercussions of preventive measures that have been implemented (1-3). Interestingly, a recent study observed that a psychiatric diagnosis prior to SARS-CoV-2 infection was significantly associated with a higher risk of COVID-19 diagnosis (4); this risk was independent of known physical risk factors and living conditions. Personality traits (i.e. relative stable patterns of feelings, thoughts and behavior) might influence disease risk by mediating health-related behaviors such as the adherence to health regulations and recommendations (e.g., social distancing or mask wearing). In line with this, studies support an inverse relationship between extroaersion and likelihood to engage in social distancing behavior at the beginning of the pandemic(5, 6).

The genetic underpinnings of psychiatric traits are known to not only show a large overlap among each other (7) but also with other diseases such as metabolic disorders(8, 9). An increased load of infections in individuals with psychiatric disorders has also been reported and may, in part, be due to shared genetic liability(10), although only few large-scale studies have tried to answer this question to date. In addition to very many other factors ranging from sex to pre-existing medical conditions and socioeconomic factors (11, 12), both common and rare genetic variants have been identified that may predispose individuals to an infection with SARS-CoV-2 or a severe course of COVID-19(13-15). A recent GWAS by the COVID-19 host genetics initiative (COVID-19 HGI(16)), identified 13 loci of genome-wide significance for the susceptibility to COVID-19, comparing subjects with self- or physician-reported COVID-19 diagnosis to the general population. Four of these loci seem to be specific to COVID-19 susceptibility rather than disease severity. The identified loci include variants in genes implicated in the innate immune response to viruses but also genomic loci harboring many genes of yet-undetermined function in the context of COVID-19.

In light of these findings, we asked whether genetic underpinnings are shared between COVID-19 susceptibility, major psychiatric disorders, and personality traits. We approached this question using both results from the largest GWAS in the respective fields and individual-level data from two observational studies of psychiatric disorders (PsyCourse) and personality traits (PsyCourse and HeiDE).

## METHODS

We performed linkage disequilibrium score regression (LDSC) (17) to calculate genetic correlations(18) between susceptibility to COVID-19 and psychiatric disorders as well as personality traits. We used summary statistics for COVID-19 susceptibility derived from a genome-wide association study (GWAS) performed by the COVID-19 Host Genetics Initiative(16) (self- or physician-reported COVID-19 diagnosis (n=87,870) vs. general population (n=2,210,804); analysis “C2” for European ancestry without 23andMe, Inc., release 6, downloaded from https://www.covid19hg.org/results/r6/, accessed July 30, 2021). For psychiatric disorders, summary data from the following GWAS were used: schizophrenia (SCZ; 33,640 cases; 43,456 controls)(19), bipolar disorder (BPD; 41,917 cases; 371,549 controls)(20), depression (as a broader phenotype closely related to major depressive disorder (MDD)) (246,363 cases; 561,190 controls)(21), and Big 5 personality traits (n=70,000 to 120,000)(22). For the details on phenotype definitions used in the GWAS, please refer to the original publications.

In a second step, individual-level data were used to calculate polygenic risk scores (PRS). In the PsyCourse Study (n=1786), consisting of individuals with major psychiatric disorders (652 SCZ, 567 BPD, 101 MDD) and controls without major psychiatric disorders (n=466), recruited throughout Germany and Austria and followed longitudinally(23), we assessed whether PRS for susceptibility to COVID-19 were associated with case status or with extraversion scores. PRS were calculated using the PRS-CS method(24), excluding the HLA region on chromosome 6. All genotyped participants of the PsyCourse Study with a diagnosis from the psychotic-to-affective spectrum as well as controls (n=1346, age [mean±SD] 47.75±13.81, 47.39% female) or for whom an extraversion score was available (n= 1190) were included in the analysis. “Case” status was defined as having a lifetime diagnosis of a severe psychiatric disorder from the spectrum of psychotic and affective disorders defined in the DSM-IV and as determined by a trained rater administering the relevant section of the SCID-I interview. The extraversion score (range: 1 to 5, mean: 3.09) was derived from a 10-item questionnaire assessing the Big 5 Personality Traits(25) (Figure). DNA samples of PsyCourse participants were genotyped on the Illumina Infinium PsychArray, and imputed using the 1000 Genomes project dataset as reference panel (for details, see(23)).

**Figure:**
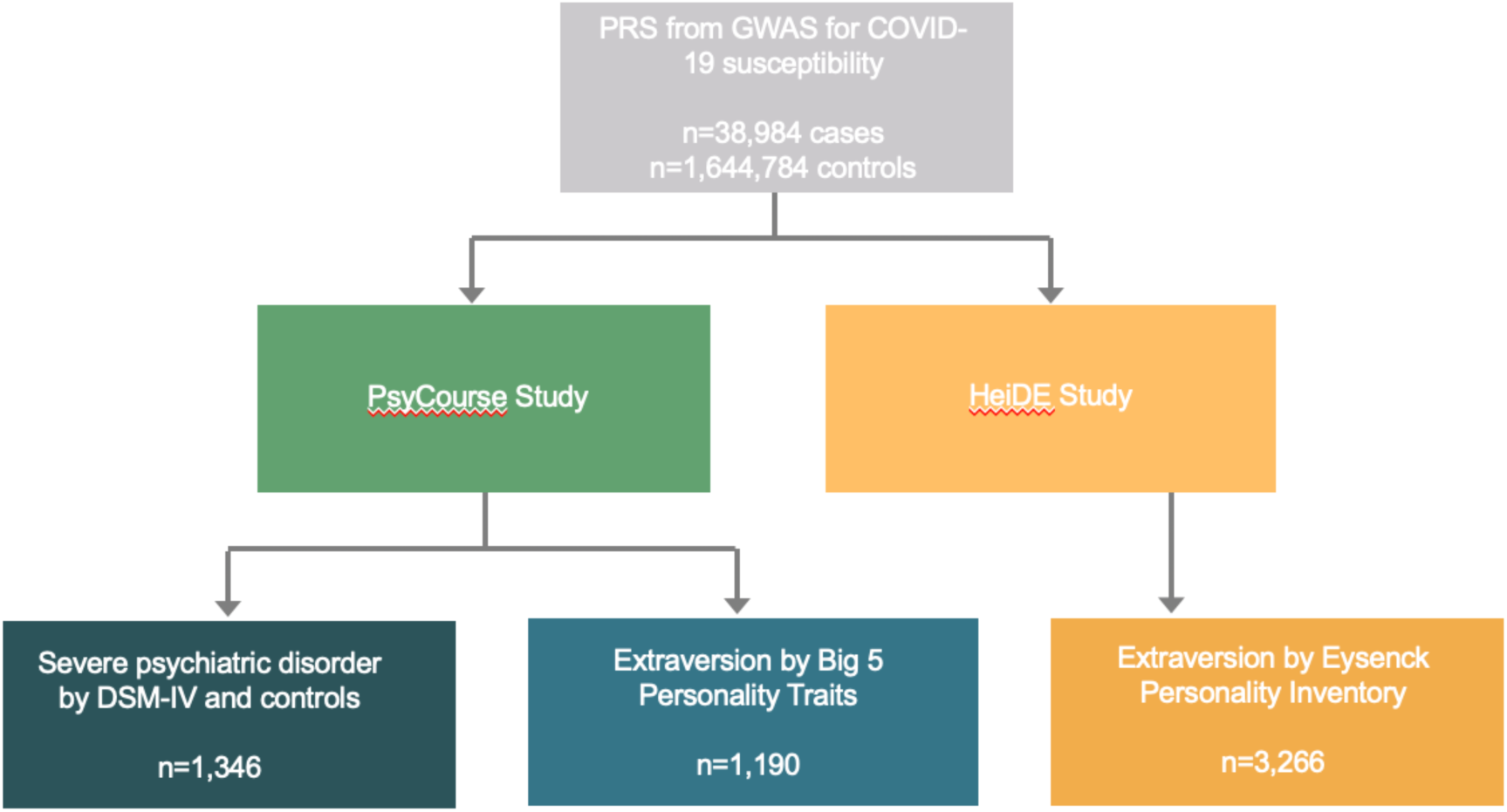
Study design for PRS analyses. Summary statistics from the to-date largest genome-wide association study on susceptibility to COVID-19 performed by the COVID-19 Host Genetics Initiative were used to generate polygenic risk scores. These were then tested for association with case/control status for severe psychiatric disorders such as schizophrenia, bipolar disorder, or major depressive disorder (PsyCourse Study only) as well as measures of extraversion (PsyCourse and HeiDE studies).

In the HeiDE study (e.g.(26)), we assessed whether extraversion scores (see below) were associated with PRS for COVID-19 susceptibility (generated using PRS-CS; n=3266, age [mean±SD] 52.78±7.06, 52.38% female). Briefly, HeiDE (“Heidelberger Langzeitstudie zu Risikofaktoren und Diagnose chronischer Erkrankungen”) is a population-based study carried out in the German city of Heidelberg and surroundings, whose initial aim was to characterize associations of personality and somatic disease. Data analyzed in this study were collected during the baseline assessment (personality traits; 1992 to 1994) and the first follow-up (DNA for genotyping; on average 8.5 years after baseline). Extraversion was measured using the Eysenck-Personality-Inventory, from which we analyzed the sum of two items closely matching the items of the Big 5 personality questionnaire used in the PsyCourse study (range: 0 to 2, mean: 1.48). DNA samples of HeiDE participants were genotyped using the Illumina Infinium PsychArray and the Infinium OmniExpress Exome Array. The combined HeiDE datasets were imputed using the 1000 Genomes phase 3 reference panel (for details, see(27)). Also see the figure for an overview of the study design.

PRS scoring and association testing using linear or ordinal regressions were implemented in PLINK (version 1.9) and R (version 4.0.3). In both studies, we regressed the respective phenotype onto age, age^2^, sex, and the first eight ancestry multi-dimensional scaling components (backward stepwise regression). The residuals of the final model were then regressed onto the PRS (PsyCourse), or the final model was compared to a model additionally containing the PRS (HeiDE). As far as we know, there is no overlap between individuals from the PsyCourse and HeiDE studies and the GWAS, whose summary statistics were used for the LDSC above.

The authors assert that all procedures contributing to this work comply with the ethical standards of the relevant national and institutional committees on human experimentation and with the Helsinki Declaration of 1975, as revised in 2008. All procedures involving human subjects were approved by the Institutional Review Board at the University of Munich (13-17), the Institutional Review Board at the Medical Faculty of the University of Heidelberg (026/2001), or the local review boards of the primary studies that the utilized summary statistics were taken from(16, 19-22, 28). Written informed consent was obtained from all study subjects.

## RESULTS

No genetic correlation was found between COVID-19 susceptibility and MDD, BPD, or SCZ risk (Table 1). When analyzing the genetic correlation between personality traits(22) and COVID-19 susceptibility, a significant positive correlation (p=1.47×10^−5^; rg=0.284) was identified for the personality trait of extraversion. No statistically significant correlation was present with any other Big-5 personality trait (Table 2). To corroborate these findings by a second, independent line of evidence using individual-level data from two independent cohorts, we turned to an assessment of PRS. In the PsyCourse Study, PRS for COVID-19 susceptibility were not significantly associated with psychiatric case status when compared to controls (p=0.474, beta=-1.132). Further, in the PsyCourse Study, no significant association between COVID-19 susceptibility PRS and extraversion as measured by the 10-item questionnaire assessing the Big-5 Personality Traits (p=0.210, beta=2.369) was found.

**Table 1:** Results from LDSC between COVID-19 susceptibility and severe psychiatric disorders. MDD=major depressive disorder, BPD=bipolar disorder, SCZ=schizophrenia, rg=genetic correlation, SE=standard error.

| Trait | MDD(21) | BPD(20) | SCZ(19) |
| --- | --- | --- | --- |
| rg | 0.072 | 0.004 | -0.037 |
| SE | 0.044 | 0.045 | 0.044 |
| p-value | 0.100 | 0.930 | 0.398 |

**Table 2:** Results from LDSC between COVID-19 susceptibility and Big 5 Personality Traits. The p-value for genetic correlation between extraversion and COVID-19 susceptibility was statistically significant. rg=genetic correlation, SE=standard error.

| **Trait** | **Agreeableness** | **Conscientiousness** | **Extraversion** | **Neuroticism** | **Openness** |
| --- | --- | --- | --- | --- | --- |
| rg | 0.177 | 0.056 | 0.284 | -0.105 | -0.095 |
| SE | 0.093 | 0.079 | 0.066 | 0.071 | 0.067 |
| p-value | 0.057 | 0.478 | 1.47E-5 | 0.142 | 0.153 |

To validate the finding for extraversion in another study setting and to mitigate any potential influence of an interaction between psychiatric disorders and personality traits in the context of COVID-19 susceptibility possibly present in the PsyCourse study, we recapitulated the extraversion analysis in the larger HeiDE study which was specifically designed to evaluate the interaction between personality traits and somatic disorders. Here, however, we also did not detect a significant association of PRS for COVID-19 susceptibility and extraversion (model comparison p=0.758, AIC 5149.51 [model with PRS] and AIC 5147.60 [model without PRS]).

## DISCUSSION

It is likely that many interdependencies exist between COVID-19 susceptibility and major psychiatric disorders or personality traits. Among these, we shed light on a potential role for shared common genetic risk factors. For major psychiatric disorders, we did not identify a significant genetic overlap that can be ascribed to common genetic variation both when assessing summary statistics of large GWAS by LDSC and when looking at PRS in individual level data, in line with emerging data in the field(16, 29, 30). With regard to personality traits, the picture is more heterogeneous with a significant signal for a positive genetic correlation between extraversion and COVID-19 susceptibility by LDSC, which needs to be explored further once larger data sets become available. However, it has to be assumed, that the genetic make-up is only one contributor in a very complex network of factors connecting extraversion to COVID-19 susceptibility.

The positive correlation identified between COVID-19 susceptibility and extraversion highlighted by the LDSC approach appears to be in line with the literature. Numerous studies performed both before and during the SARS-CoV-2 pandemic have demonstrated the effect of personality determinants on health behavior and outcomes (e.g.(5, 31, 32)). For example, it was shown that narcissistic tendencies coincide with decreased perceived susceptibility to infection with SARS-CoV-2(33) while, at least for neuroticism, not genetic overlap was found(29). Intuitively, less extroverted individuals may find social distancing during the pandemic easier than extroverted individuals and may, therefore, be more compliant with social distancing rules and at an overall decreased risk for COVID-19(5, 6). There is even evidence of a bidirectionality of this phenomenon—the general risk for infectious diseases in a given region may, in part, influence personality traits at population level such that lower mean levels of extraversion are reported in regions with higher prevalence of infectious diseases(34). One possible reason for this could be that in regions where ever-present infectious diseases present a comparatively large thread to health and well-being, less extraversion is present at population level either because people have adapted their behavior or because of potential selective pressure. Yet, it is likely that many interdependencies exist between COVID-19 susceptibility and personality traits or major psychiatric disorders and we investigated only shared common genetic risk factors.

Although all included GWAS are the currently largest in the respective fields, sample sizes may still not be large enough to confidently detect genetic correlations in settings with many natural confounders such as levels of exposure to the virus or socioeconomic differences, to name only a few. Also, different instruments where used to evaluate personality traits in PsyCourse and HeiDE and the study populations (individuals with severe psychiatric disorders and controls vs. the general population) were different, possibly contributing to the observed heterogeneity. While LDSC represents a powerful tool to assess genetic correlations, other methods to quantify polygenic overlap irrespective of genetic correlations also exist (e.g.,(35)) and could be used to explore potential shared genetic underpinnings in even greater depth but are beyond the scope of this study. An additional limitation lies in the fact that no direct risk assessment was possible for the individuals with individual-level data on major psychiatric disorders and personality traits since no COVID-19 phenotypes were available. Lastly, we are unable to fully exclude sample overlap especially on the side of the controls used in the included GWAS. However, LDSC results should be robust to both of these overlaps(18).

Hypothetically, it is possible that—for example—only a small subset of common genetic risk factors in a given pathway relevant to major psychiatric disorders or personality traits is associated with COVID-19 susceptibility. Although we cannot fully exclude all such effects, our data suggest that non-genetic factors play important roles in the interplay between personality traits and COVID-19. A direct genetic overlap is unlikely to contribute to the increased, but yet-unexplained COVID-19 risk seen in individuals with a psychiatric diagnosis prior to SARS-CoV-2 infection(4) but a shared genetic risk could still be mediated by intermediate phenotypes such as, for example, lower socio-economic status or educational attainment in those with severe psychotic disorders. As a consequence, an even greater focus should be placed on psychosocial interventions, ensuring the best treatment for individuals with severe psychiatric disorders as well as targeted measures of prevention and psychoeducation for individuals with personality determinants that place them at an increased pandemic-related risk for health and well-being.

## Data Availability

Individual level data from the PsyCourse Study (www.psycourse.de) are freely available to all researchers upon submission of a proposal and granting of access rights by the study's data access committee (please see description of the application procedure on the study's website). Individual level data from the HeiDE study are available upon request to the corresponding author.

## ACKNOWLEDGEMENTS & FUNDING

We are extremely grateful to all members of the COVID-19 Host Genetics Initiative for rapidly sharing data in an open-science fashion and to all study participants of all the studies included in the COVID-19 Host Genetics Initiative (https://www.covid19hg.org/acknowledgements/), the PsyCourse Study (http://www.psycourse.de), and the HeiDE Study without whose contributions this work would not have been possible. Further, our work also depended upon the sharing of summary statistics from the large GWAS on SCZ, BPD, and MDD performed by the Psychiatric Genomics Consortium (PGC) and as well as the authors and participants of the Big 5 GWAS performed by Lo et al. We would like to thank the research participants and employees of 23andMe for making this work possible. MMN and ECS are members of the German COVID-19 Omics Initiative (DeCOI). MMN is member of the DFG-funded Excellence Cluster ImmunoSensation2 (EXC 2151 – 390873048). This study was supported by ERA-NET NEURON grants “EMBED” (01EW1904 to MR), and “SynSchiz—Linking synaptic dysfunction to disease mechanisms in schizophrenia—a multilevel investigation” (01EW1810 to MR). TGS and PF are supported by the German Research Foundation (Deutsche Forschungsgemeinschaft; DFG) within the framework of the projects www.kfo241.de and www.PsyCourse.de (SCHU 1603/4-1, 5-1, 7-1; FA241/16-1), ECS is also supported by the DFG (SCHU 2419/2-1) and through the Munich Clinician Scientist Program (MCSP). The genotyping was in part funded by the German Federal Ministry of Education and Research (BMBF) through the Integrated Network IntegraMent (Integrated Understanding of Causes and Mechanisms in Mental Disorders), under the auspices of the e:Med Program with grants awarded to TGS (01ZX1614K), MR (01ZX1614G), and MMN (01ZX1614A). TGS received additional support from the German Federal Ministry of Education and Research (BMBF) within the framework of the BipoLife network and the Dr. Lisa Oehler Foundation, Kassel (Germany). OAA received support from Research Council of Norway (#223273, #*283798*, #248980) and NordForsk (#105668). We thank Stefan Herms for his support in the technical provision of genotype data.

## AUTHOR CONTRIBUTIONS

Formulation of research question: UH, FS, MR, TGS, ECS

Study design: UH, FS, TS, AL, MA, OAA, CHC, PF, MR, TGS, ECS

Study conduction: UH, TV, FS, SKS, DRE, FKS, JLK, MH, KG, ALC, MB, HAS, KA, TS, AL, MA, JZ, JW, MVH, CS, MS, ER, JR, CK, GJ, FUL, MJ, CF, AJFa, DED, UD, BTB, VA, IGA, MMN, SHH, AJFo, FD, SHW, OAA, CHC, PF, MR, TGS, ECS

Data analysis: UH, FS, SP, MOK, TFMA, EP, JF, OAA, CHC, ECS

Manuscript writing: UH, FS, ECS

Critical revision of manuscript: all authors

## DATA AVAILABILITY

The data that support the findings in this study are available from the corresponding author, ECS, upon reasonable request. The relevant summary statistics from the GWAS used in the analyses are available from the authors of the primary studies(16, 19-22, 28). Interested researchers can also apply for the used as well as additional data for the PsyCourse Study via http://www.psycourse.de/openscience-en.html.

## REFERENCES

1. Pan KY, Kok AAL, Eikelenboom M, Horsfall M, Jorg F, Luteijn RA, et al. The mental health impact of the COVID-19 pandemic on people with and without depressive, anxiety, or obsessive-compulsive disorders: a longitudinal study of three Dutch case-control cohorts. Lancet Psychiatry. 2021;8(2):121–9.

2. Pfefferbaum B, North CS. Mental Health and the Covid-19 Pandemic. N Engl J Med. 2020;383(6):510–2.

3. Wang Y, Shi L, Que J, Lu Q, Liu L, Lu Z, et al. The impact of quarantine on mental health status among general population in China during the COVID-19 pandemic. Mol Psychiatry. 2021.

4. Taquet M, Luciano S, Geddes JR, Harrison PJ. Bidirectional associations between COVID-19 and psychiatric disorder: retrospective cohort studies of 62 354 COVID-19 cases in the USA. Lancet Psychiatry. 2021;8(2):130–40.

5. Carvalho LF, Pianowski G, Goncalves AP. Personality differences and COVID-19: are extroversion and conscientiousness personality traits associated with engagement with containment measures? Trends Psychiatry Psychother. 2020;42(2):179–84.

6. Gotz FM, Gvirtz A, Galinsky AD, Jachimowicz JM. How personality and policy predict pandemic behavior: Understanding sheltering-in-place in 55 countries at the onset of COVID-19. Am Psychol. 2021;76(1):39–49.

7. Cross-Disorder Group of the Psychiatric Genomics Consortium. Electronic address pmhe, Cross-Disorder Group of the Psychiatric Genomics C. Genomic Relationships, Novel Loci, and Pleiotropic Mechanisms across Eight Psychiatric Disorders. Cell. 2019;179(7):1469–82 e11.

8. Bahrami S, Steen NE, Shadrin A, O’Connell K, Frei O, Bettella F, et al. Shared Genetic Loci Between Body Mass Index and Major Psychiatric Disorders: A Genome-wide Association Study. JAMA Psychiatry. 2020;77(5):503–12.

9. Hubel C, Gaspar HA, Coleman JRI, Hanscombe KB, Purves K, Prokopenko I, et al. Genetic correlations of psychiatric traits with body composition and glycemic traits are sex-and age-dependent. Nat Commun. 2019;10(1):5765.

10. Nudel R, Wang Y, Appadurai V, Schork AJ, Buil A, Agerbo E, et al. A large-scale genomic investigation of susceptibility to infection and its association with mental disorders in the Danish population. Transl Psychiatry. 2019;9(1):283.

11. de Lusignan S, Dorward J, Correa A, Jones N, Akinyemi O, Amirthalingam G, et al. Risk factors for SARS-CoV-2 among patients in the Oxford Royal College of General Practitioners Research and Surveillance Centre primary care network: a cross-sectional study. Lancet Infect Dis. 2020;20(9):1034–42.

12. Docherty AB, Harrison EM, Green CA, Hardwick HE, Pius R, Norman L, et al. Features of 20 133 UK patients in hospital with covid-19 using the ISARIC WHO Clinical Characterisation Protocol: prospective observational cohort study. BMJ. 2020;369:m1985.

13. Pairo-Castineira E, Clohisey S, Klaric L, Bretherick AD, Rawlik K, Pasko D, et al. Genetic mechanisms of critical illness in Covid-19. Nature. 2020.

14. Severe Covid GG, Ellinghaus D, Degenhardt F, Bujanda L, Buti M, Albillos A, et al. Genomewide Association Study of Severe Covid-19 with Respiratory Failure. N Engl J Med. 2020;383(16):1522–34.

15. Zhang Q, Bastard P, Liu Z, Le Pen J, Moncada-Velez M, Chen J, et al. Inborn errors of type I IFN immunity in patients with life-threatening COVID-19. Science. 2020;370(6515).

16. Initiative C-HG. Mapping the human genetic architecture of COVID-19. Nature. 2021.

17. Bulik-Sullivan BK, Loh PR, Finucane HK, Ripke S, Yang J, Schizophrenia Working Group of the Psychiatric Genomics C, et al. LD Score regression distinguishes confounding from polygenicity in genome-wide association studies. Nat Genet. 2015;47(3):291–5.

18. Bulik-Sullivan B, Finucane HK, Anttila V, Gusev A, Day FR, Loh PR, et al. An atlas of genetic correlations across human diseases and traits. Nat Genet. 2015;47(11):1236–41.

19. Schizophrenia Working Group of the Psychiatric Genomics C. Biological insights from 108 schizophrenia-associated genetic loci. Nature. 2014;511(7510):421–7.

20. Mullins N, Forstner AJ, O’Connell KS, Coombes B, Coleman JRI, Qiao Z, et al. Genome-wide association study of more than 40,000 bipolar disorder cases provides new insights into the underlying biology. Nat Genet. 2021.

21. Howard DM, Adams MJ, Clarke TK, Hafferty JD, Gibson J, Shirali M, et al. Genome-wide meta-analysis of depression identifies 102 independent variants and highlights the importance of the prefrontal brain regions. Nat Neurosci. 2019;22(3):343–52.

22. Lo MT, Hinds DA, Tung JY, Franz C, Fan CC, Wang Y, et al. Genome-wide analyses for personality traits identify six genomic loci and show correlations with psychiatric disorders. Nat Genet. 2017;49(1):152–6.

23. Budde M, Anderson-Schmidt H, Gade K, Reich-Erkelenz D, Adorjan K, Kalman JL, et al. A longitudinal approach to biological psychiatric research: The PsyCourse study. Am J Med Genet B Neuropsychiatr Genet. 2019;180(2):89–102.

24. Ge T, Chen CY, Ni Y, Feng YA, Smoller JW. Polygenic prediction via Bayesian regression and continuous shrinkage priors. Nat Commun. 2019;10(1):1776.

25. Rammstedt BaJ, O.P. Measuring personality in one minute or less: A 10-item short version of the Big Five Inventory in English and German. Journal of Research in Personality. 2007;41(1):203–12.

26. Sturmer T, Hasselbach P, Amelang M. Personality, lifestyle, and risk of cardiovascular disease and cancer: follow-up of population based cohort. BMJ. 2006;332(7554):1359.

27. Heilbronner U, Papiol S, Budde M, Andlauer TFM, Strohmaier J, Streit F, et al. “The Heidelberg Five” personality dimensions: Genome-wide associations, polygenic risk for neuroticism, and psychopathology 20 years after assessment. Am J Med Genet B Neuropsychiatr Genet. 2021;186(2):77–89.

28. Wray NR, Ripke S, Mattheisen M, Trzaskowski M, Byrne EM, Abdellaoui A, et al. Genome-wide association analyses identify 44 risk variants and refine the genetic architecture of major depression. Nat Genet. 2018;50(5):668–81.

29. Chang XL, Yun; Nguyen, Kenny; Qu, Huiqi; Liu, Yichuan; Glessner, Joseph; Sleiman, Patrick M. A.; Hakonarson, Hakon. Genetic correlations between COVID-19 and a variety of traits and diseases. 2021.

30. Luykx JJ, Lin BD. Are psychiatric disorders risk factors for COVID-19 susceptibility and severity? a two-sample, bidirectional, univariable, and multivariable Mendelian Randomization study. Transl Psychiatry. 2021;11(1):210.

31. Eichenberg C, Grossfurthner M, Andrich J, Hubner L, Kietaibl S, Holocher-Benetka S. The Relationship Between the Implementation of Statutory Preventative Measures, Perceived Susceptibility of COVID-19, and Personality Traits in the Initial Stage of Corona-Related Lockdown: A German and Austrian Population Online Survey. Front Psychiatry. 2021;12:596281.

32. Friedman HS, Kern ML. Personality, well-being, and health. Annu Rev Psychol. 2014;65:719–42.

33. Venema TAG, Pfattheicher S. Perceived susceptibility to COVID-19 infection and narcissistic traits. Pers Individ Dif. 2021;175:110696.

34. Schaller M, Murray DR. Pathogens, personality, and culture: disease prevalence predicts worldwide variability in sociosexuality, extraversion, and openness to experience. J Pers Soc Psychol. 2008;95(1):212–21.

35. Frei O, Holland D, Smeland OB, Shadrin AA, Fan CC, Maeland S, et al. Bivariate causal mixture model quantifies polygenic overlap between complex traits beyond genetic correlation. Nat Commun. 2019;10(1):2417.

